# How Previous Epidemics Enable Timelier COVID-19 Responses: A Cross-Sectional Study Using Organizational Memory Theory

**DOI:** 10.1101/2020.06.23.20138479

**Authors:** Sian Hsiang-Te Tsuei

## Abstract

**Introduction:** There has been little systematic exploration of what affects timeliness of epidemic response, despite the potential for earlier responses to be more effective. Speculations have circulated that exposure to major epidemics helped health systems respond more quickly to COVID-19. This study leverages organizational memory theory to test whether health systems with any, more severe, more recent exposure to major epidemics enacted timelier COVID-19 policy responses.

**Methods:** A dataset was constructed cataloguing 846 policies across 178 health systems in total, 37 of which had major epidemics within the last twenty years. Hypothesis testing used OLS regressions with WHO region fixed effects, controlling for several health system expenditure and political variables.

**Results:** Results show that exposure to any major epidemics was significantly associated with providing earlier response in any category or for surveillance / response, distancing, and international travel policies when tested alone or with total number of cases. The effect was about six to ten days earlier response. The significance was largely nullified with additional independent variables. Both total cases and years since previous epidemics showed no statistical significance.

**Conclusion:** This study suggests that health systems may learn from past major epidemics. Policymakers ought to institutionalize lessons from COVID-19. Future studies can examine specific generalizable lessons and whether timelier responses correlated with lower health and economic impacts.

**What is already known about this subject?:** There has been little systematic exploration into what affects the timeliness of response to epidemics. For COVID-19, there has been speculation that previous exposure to major epidemics may have spurred timelier responses.

**What are the new findings?:** Applying organizational memory theory, this study identified policy response timeliness to COVID-19 was significantly associated with any past exposure to major epidemics within the last 20 years.

**What are the recommendations for policy and practice?:** The fact that any exposure to past epidemics is significantly associated with timelier responses suggest that health systems do learn from past mistakes. Institutionalizing the learnings through more permanent policies. Once COVID-19 fades, health systems may wish to revitalize the organizational memory through various exercises.

## Introduction

### Background

The drastic consequences of COVID-19 highlight the need to identify tactics that could have mitigated these outcomes. Earlier interventions are particularly worthwhile, as such policies might use simpler techniques such as border control or isolated case finding followed by quarantine. In contrast, interventions once a disease has established a foothold may require expensive life-supporting therapies or aggressive public health measures that might cause significant socioeconomic disruptions. Later interventions may be also less effective. Asymptomatic transmission as is the case for COVID-19 may elude effective containment strategies unless implemented universally, leading to exponential case growth that overwhelm the health system. Therefore, it is particularly important to examine how the pandemic could have been limited to earlier stages.

There has been report of improving timeliness of disease outbreak recognition [1]. However, there is little rigorous exploration on what drives timeliness of policy response to epidemics. Kandel et al. examined in 2020 the capacity for 182 countries to respond to public health emergencies [2]. They constructed operational readiness index based on select metrics from the domains of a) technical abilities, including the ability to conduct surveillance, contact tracing, and infection prevention; b) governance capacity, marked by capacity for multisectoral collaboration, emergency risk communication; and c) resource availability, defined in terms of finances, and human resources. However, these indices were constructed neither systematically nor empirically.

Other attempts to explore epidemic response timeliness have typically drawn on sporadic *post hoc* reviews or case studies. For example, the United States (US) Center for Disease Control (CDC) documented that it responded quickly within the month that the Middle East Respiratory Syndrome Coronavirus (MERS-CoV) case was reported to the online Program for Monitoring Emerging Diseases. It mobilized its personnel to begin collecting more information, quickly developed briefings for staff members and the general public, and posting travel updates [3].

The literature has typically shied away from rigorous frameworks to examine how timely response to epidemics could be enacted. One study, however, did attempt such a methodology. Hanvoravongchai et al. examined the six Asian health systems’ preparedness for an influenza pandemic [4]. They concluded that more prepared health systems had pandemic response integrated into national disaster preparedness framework, lower public health official turnovers, previous pandemic exposures, and more political emphasis on preparedness. Nevertheless, their work was focused on generating hypotheses rather than testing hypotheses, and as such, there is a dearth of literature that systematically tests a developed hypothesis on what affects timeliness of epidemic response.

### Objectives

This study aims to explore why health systems differ in their timeliness of issuing policy responses to COVID-19 before the epidemic took hold within their health system. Exploring what drives timelier response may point future health system researchers in fruitful directions.

The WHO has defined health system explicitly to capture “all the activities whose primary purpose is to promote, restore or maintain health [5]”. Among the various actors within health system, this study specifically focuses on the executive branch of the government. Their attempt to minimize COVID-19 burden through prevention is captured within the definition of health system. The focus on such an organizational consideration is particularly worthwhile, since it is traditionally more opaque and de-emphasized particularly among low- and middle-income countries [6]. This is unfortunate because it is the dynamics and bureaucratic abilities within the guiding government bodies that affect the policy development. Specifically, the dynamics within the executive branch of government determine policy responses, and this branch is particularly relevant for epidemic responses. First, they can respond most swiftly in an emergency. The legislative branch often requires passing several readings of the law and the judicial branch is responsible for *ex post* interpretation of laws. Second, the executive branch drives the responses of subsidiary government organizations, providing targets and support for organizations such as the Center for Disease Control. Here, the executive branch is considered as a single decision-making unit, experiencing the relevant dynamics together, since the relevant parties within this unit must reach a consensus on the relevant policies.

The popular media claims that the countries with more serious exposures to SARS or MERS-CoV responded more appropriately to COVID-19 [7,8]. The intuition is that the health systems faced with infections with potentially high risk of case fatality rate in the past have learned from their past experiences, and can therefore cope better, be it more promptly or more effectively. In this case, the diseases identified have been found to have case fatality rate of greater than 1%.

Such intuitive reasoning resonates with both organizational memory theory and institutional memory theory. *Organizational memory* is defined as the “stored information from an organization’s history that can be brought to bear on present decisions.”[9], and institutional memory is defined similarly as organizational elites’ “shared knowledge…about past crisis management” [10]. Given the richer development of the organizational memory theory, however, I favour the organizational memory term for the rest of the paper. In Walsh and Ungson’s seminal review of organizational memory, they describe that relevant information about a problem and associated decision is encoded within an organization via several methods, such as individuals’ memories, organizational procedures, and organizational cultures [9]. When a similar problem surfaces again, the organization retrieves the stored information. Assuming that the executive branch personnel aim for timelier responses, this therefore leads to the first hypothesis:

**Hypothesis 1**: Having at least one case of a disease that is highly threatening within the last twenty years is associated with more timely response.

The duration of the lessons may be non-permanent depending on the way the lessons were encoded. If encoded into policies, the lessons may last as long as the institution itself. However, Walsh and Ungson suggest that much of the organizational memory is encoded within the individuals’ memories [9], rendering the lessons impermanent. As more individuals exposed to the problem retire, the relevant lessons and expertise fades. With more recent exposures, the memory of the relevant procedures allows more appropriate response. This therefore generates the second hypothesis:

**Hypothesis 2**: The more time elapsed since the last major epidemic in a health system, the slower the response in the health system.

Psychology has shown also that the depth of memory correlates with the intensity of the emotions during the event [11,12]. The stronger the emotion, the stronger the activation of emotional processing center amygdala, leading to more lasting encoding of the memories in the hippocampus [13]. The more severe the previous epidemic, the more likely that the involved individuals would retain the lessons for prolonged period. It follows then that the more serious the last epidemic the executive branch faced, the deeper the episodic memory.

**Hypothesis 3**: The more cases a health system dealt with in past major epidemics, the timelier the policy responses in the health system.

## Methods

### Study design

This study uses a cross-sectional regression to assess the association between exposure to major epidemics previously and timeliness of COVID-19 response.

Major epidemics were included if they: occurred within the last 20 years (i.e., years 2000 – 2020), were documented within the WHO Disease Outbreak Network, had case fatality rate > 1%, affected multiple health systems, and affected more than 500 cases in total. Diseases were excluded if they had vaccines available at the time of the outbreak, were vector-borne diseases, or were COVID-19. This process resulted in the following diseases: SARS, H1N1, Ebola, and MERS. However, given that estimates for H1N1’s case fatality rate is generally lower and more variable, separate analyses were conducted without H1N1.Health systems were considered to have been affected by these diseases if they had >100 cases of the disease during the outbreak (i.e., not relying on retrospective modeling parameters). All countries listed in World Bank database were considered. Health systems were parsed out from countries list if they have unique health system operations1. This resulted in a total of 177 health systems.

The number of policies considered were the first policies of each of the first five WHO categories. The total number of policies in the dataset were 846 policies, made up of 82 individual measures, 91 environmental measures, 177 surveillance and response measures, 177 social and physical distancing measures, and 188 international travel measures.

### Dataset and variables

The timeliness of response was the difference in days between the policy issuance and January 5^th^, 2020, when WHO first announced the pneumonia of unknown significance. The policies are grouped according to the WHO Codes, separated into individual measures, environmental measures, surveillance and response measures, social and physical distancing measures, and international travel measures. One category of policy was coded separately to capture the first of all policies within the health system. Each health system therefore would generate one unique first policy overall, and one first policy per WHO category. For completeness, the timeliness of response was also calculated with regards to December 31^st^, 2019, which was when China first notified its WHO division with regards to pneumonia of unknown significance. This was examined as well, given that some health systems such as Taiwan reacted on that day.

Exposure to any major epidemics accounts for exposure to any of the included diseases. Total number of cases tabulates the total number of cases resulting from the included diseases. Years since epidemics was calculated in terms of the number of years between 2020 and the last year the disease occurred in the health system. The dataset also included several variables that may affect executive governments’ decision process and confound the relationship between policy timeliness and organizational memory. Covariates include corruption, fiscal spending, and executive government’s political preferences. Continent-specific cultures are proxied using WHO regions. Health care spending priority is proxied using total health expenditure as a proportion of total government revenue.

The covariates were chosen based on theory. Relying on a deterministic framework is particularly useful given the complexity of the health system and the potential for numerous interconnected pathways. The choices leaned specifically on the renowned control knob framework [14], which describes the factors that determine the various outcomes of interest in a health system. The major control knobs include financing, the amount of money available for health-related activities; payment, the method of reimbursement for health-system labour; organization, the characteristics of health system organization; regulation, the various policies available for guiding health system actors; and persuasion, which include promotional activities that affect people’s preferences.

The main covariates in this study mainly relates to financing and organization. Financing capacity is captured by the variables on GDP per capita and the proportion of GDP spent on health. Organizational characteristics considered include political leanings and the extent of corruption. This study does not control for payment, regulation, and persuasion control knobs, and the implications are addressed in the *Bias* section of the study.

Another covariate controlled for the date of the first case in the health system. The timeliness of policy responses likely was affected by the timing of the first case, and if the entry of the first case correlated with previous exposure to any of the three independent variables of interest, this could result in confounding.

The list of datasets and variables are provided in table 1 and table 2, respectively.

**Table 1:** List of datasets

| **Dataset Name** | **Content** | **Source(s)** |
| --- | --- | --- |
| Past Epidemics exposure | Number of cases, deaths, and years affected by SARS, H1N1, MERS, Ebola<br>Note: Saudi Arabia's MERS burdens were hand coded based on review of the monthly reports of Disease outbreak News[18]. Data available upon request. Seychelles' international travel ban was modified to February 3<sup>rd</sup>, 2020 [19]. | [18,20–24] |
| COVID-19 cases and deaths | Day-by-day number of cases and deaths of COVID-19 | [25] |
| Public Health and Social Measures | Number, type, and content of policy measures for COVID-19 as classified by WHO and up-to-date as of May 14<sup>th</sup>, 2020 | [26] |
| Corruption Perception Index | Population's perception of corruption | [27] |
| GDP per capita | GDP per capita in current US Dollars in 2018<br>Note: Taiwan's GDP per capita was supplemented[28]. | [29] |
| Health priority | Health expenditure as a percentage of all GDP in 2018 | [30] |
| Population size | Number of people in the population in 2018<br>Note: Taiwan's population was supplemented [31]. | [32] |

**Table 2:** List of variables

| Variable name | Operational definition | Type |
| --- | --- | --- |
| **Key Dependent Variable** |  |  |
| *Firstpol* | Delay of first policy of each WHO category relative to Jan 5, 2020<sup>3</sup> (i.e., when WHO first announced pneumonia of unknown significance) | Continuous |
| **Key independent variables** |  |  |
| *Anyepi* | History of exposure to major epidemics in last 20 years | Binary (Yes = 1; No = 0) |
| *Totcase* | Total number of cases from major epidemics within the last 20 years. (Note: health systems with less than 100 cases of an outbreak were coded as having 0 cases.) | Continuous |
| *Yr_since* | Number of years elapsed since the last major epidemic. Health systems without major epidemics were coded as NA. | Continuous |
| **Covariate Variables** |  |  |
| *Corrupt* | Corruption index in 2019 | Continuous |
| *GDPcap* | GDP per capita in 2018 | Continuous |
| *Hlthpri* | Health care priority, as defined by total health expenditure as a fraction of total government revenue in 2018 | Continuous |
| *Execrlc* | Executive asbranch's political leaning towards right (1), center (2), left (3), no information (0), or no executive branch (NA) in 2017 | Categorical |
| *WHO_Region* | Regions of world as denoted by WHO classification in 2020 | Categorical |

<sup>3</sup> For completeness, the variable was also defined relative to December 31, 2019, which was when China reported pneumonia of unknown significance to the WHO.

### Statistical methods

The estimations relied on variations of equation 1.

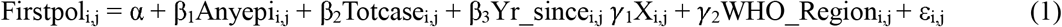

Equation 1 tests whether the first policy relative to Jan 5, 2020 for health system *i* in WHO region *j* is associated with exposure to any epidemics (β_1_), total cases (β_2_), and years since last major epidemics (β_3_) while controlling for the vector of health system covariates *X*^*2*^. The *firstpol* variable indicate the health system’s very first policy, as well as the health system’s first policy within each WHO category.

The equation varied by testing all permutations of the three independent variables. The permutations included a single variable, combinations of two variables, and all three variables together. The equation also used WHO Region fixed effects. Standard errors were not clustered to minimize false rejections of the null hypothesis.

Ideally, all three key independent variables can be tested together in one equation, but the collinearity between *Anyepi* and *Yr_since* precludes simultaneous testing.

β_1_ to β_3_ are expected to be negative, suggesting that any exposure to epidemics or higher total cases from epidemics. β_4_ is expected to be positive to indicate that longer time elapsed since last major epidemic is associated with longer policy delay.

### Bias

Residual confounding may arise in three separate places. First, the lack of control for other control knobs may be the most potent cause of residual confounding. While this was in part due to lack of available data, I suggest some reasons why the potential bias may be minimal. The payment methods for the executive branch of the government is typically salary-based, given that this is usually how governments pay their employees. The understandably uniform method of payment lowers the potential for bias. Regulation and persuasion with regards to COVID-19 are important for affecting the timeliness of response, but they were understandably missing at the outset of the pandemic. In fact, the timeliness of the policy implementation for regulations and persuasions are the matter of the study. Second, some of the indices were available only for 2017 or 2018. The regression dropped health systems for which there were missing covariates. This differential attrition typically involves underdeveloped countries. Because these countries could have responded either more quickly or more slowly to COVID-19, it is difficult to predict the direction of bias. Third, the study controls for the date of first case in the health system but does not control for when the first case arrived in a neighbouring health system or region. This may also be a form of residual confounding.

### Patient and public involvement

Patients or the public were not involved in the design, or conduct, or reporting, or dissemination plans of the research.

## Results

### Descriptive data

On average, the delay of first policy relative to the first WHO announcement of COVID-19 was approximately 54 days. About 25% of the dataset included policies from health systems exposed to any history of epidemics, translating to 41 health systems having been exposed to any major epidemics within the last 20 years. The epidemics affected on average about 243 cases. Among health systems exposed to major epidemics, the average was about 9 years since last epidemic. The values of these variables were largely the same when the dataset excluded H1N1 as a major epidemic.

The corruption index was 44, suggesting that on balance, the governments may be more corrupt. The GDP per capita was USD 16,250, suggesting that the average is skewed towards high income countries. The level of expenditure for health priority spending appears to be closer towards low- and middle-income countries’ spending than high income countries’. More of the systems exhibited either more rightist or leftist governments, with somewhat lower representation from the South-East Asia Region; and WPRO, Western Pacific Region. The descriptive data is summarized in table 3.

**Table 3:**
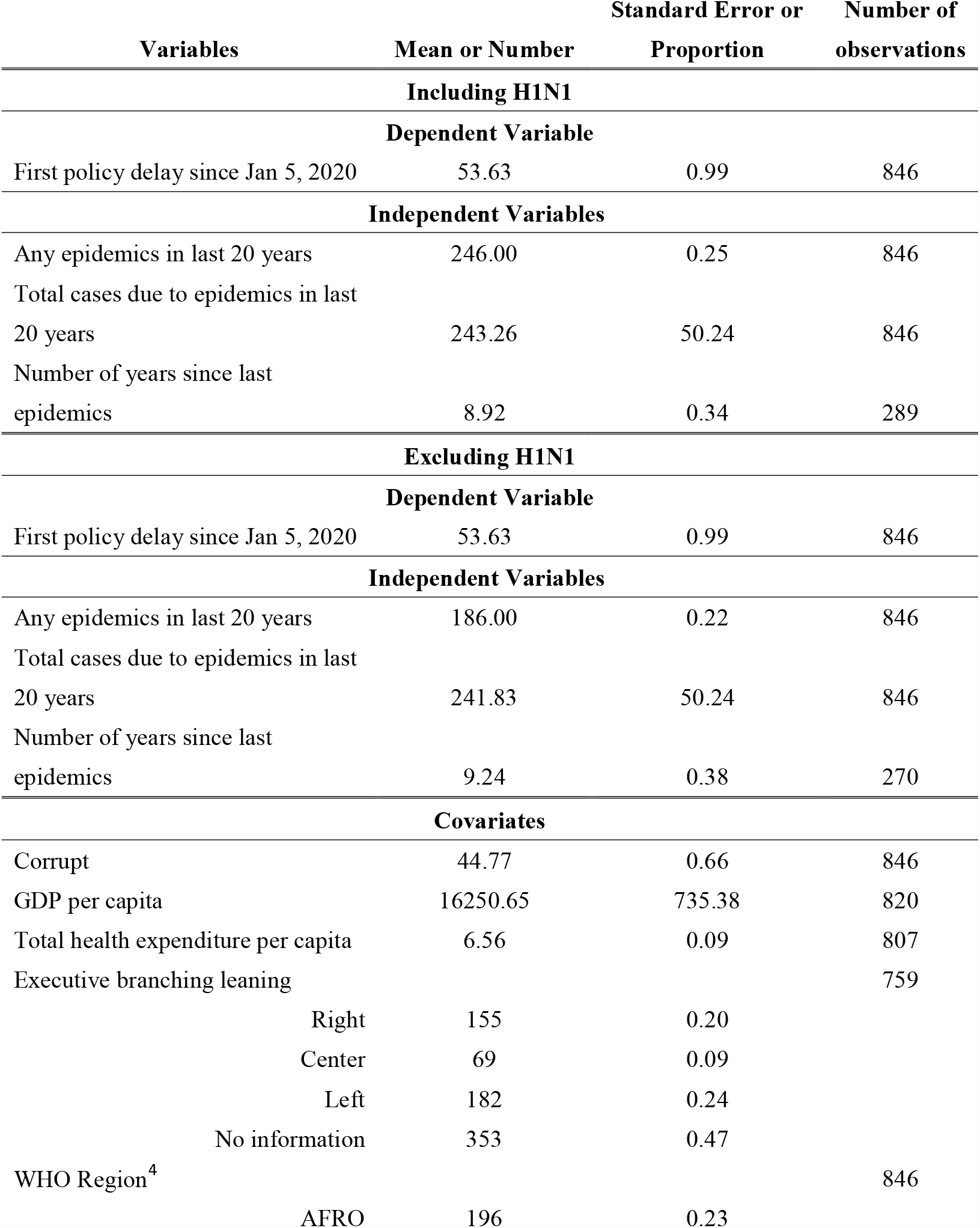
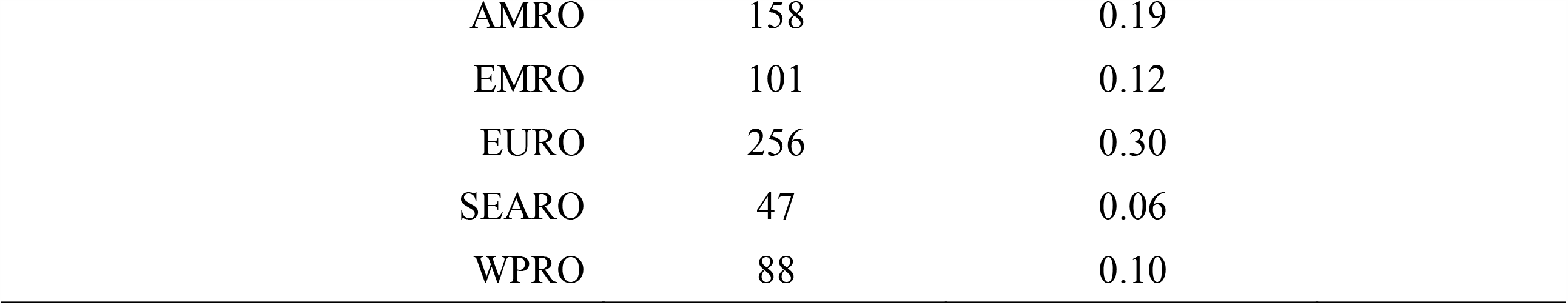
Descriptive Statistics

| **Variables** | **Mean or Number** | **Standard Error or Proportion** | **Number of observations** |
| --- | --- | --- | --- |
| **Including H1N1** |  |  |  |
| **Dependent Variable** |  |  |  |
| First policy delay since Jan 5, 2020 | 53.63 | 0.99 | 846 |
| **Independent Variables** |  |  |  |
| Any epidemics in last 20 years | 246.00 | 0.25 | 846 |
| Total cases due to epidemics in last 20 years | 243.26 | 50.24 | 846 |
| Number of years since last epidemics | 8.92 | 0.34 | 289 |
| **Excluding H1N1** |  |  |  |
| **Dependent Variable** |  |  |  |
| First policy delay since Jan 5, 2020 | 53.63 | 0.99 | 846 |
| **Independent Variables** |  |  |  |
| Any epidemics in last 20 years | 186.00 | 0.22 | 846 |
| Total cases due to epidemics in last 20 years | 241.83 | 50.24 | 846 |
| Number of years since last epidemics | 9.24 | 0.38 | 270 |
| **Covariates** |  |  |  |
| Corrupt | 44.77 | 0.66 | 846 |
| GDP per capita | 16250.65 | 735.38 | 820 |
| Total health expenditure per capita | 6.56 | 0.09 | 807 |
| Executive branching leaning |  |  | 759 |
| Right | 155 | 0.20 |  |
| Center | 69 | 0.09 |  |
| Left | 182 | 0.24 |  |
| No information | 353 | 0.47 |  |
| WHO Region<sup>4</sup> |  |  | 846 |
| AFRO | 196 | 0.23 |  |

<sup>4</sup> The WHO regions are defined as follows: AFRO, African region; AMRO, Americas; EMRO, Eastern Mediterranean; EURO, European Region; SEARO, South-East Asia Region; and WPRO, Western Pacific Region.

|  |  |  |
| --- | --- | --- |
| AMRO | 158 | 0.19 |
| EMRO | 101 | 0.12 |
| EURO | 256 | 0.30 |
| SEARO | 47 | 0.06 |
| WPRO | 88 | 0.10 |

In aggregate, the policies mostly centered around February and mid-March. When broken down by categories, the bulk of the international travel and surveillance / response policies occurred first, occurring around February. The distancing measures then appeared, concentrated mostly around mid-March. Subsequently, the individual and environmental policies occurred, concentrating mostly around April to May. The policy distributions are summarized in figure 1.

**Figure 1e:**
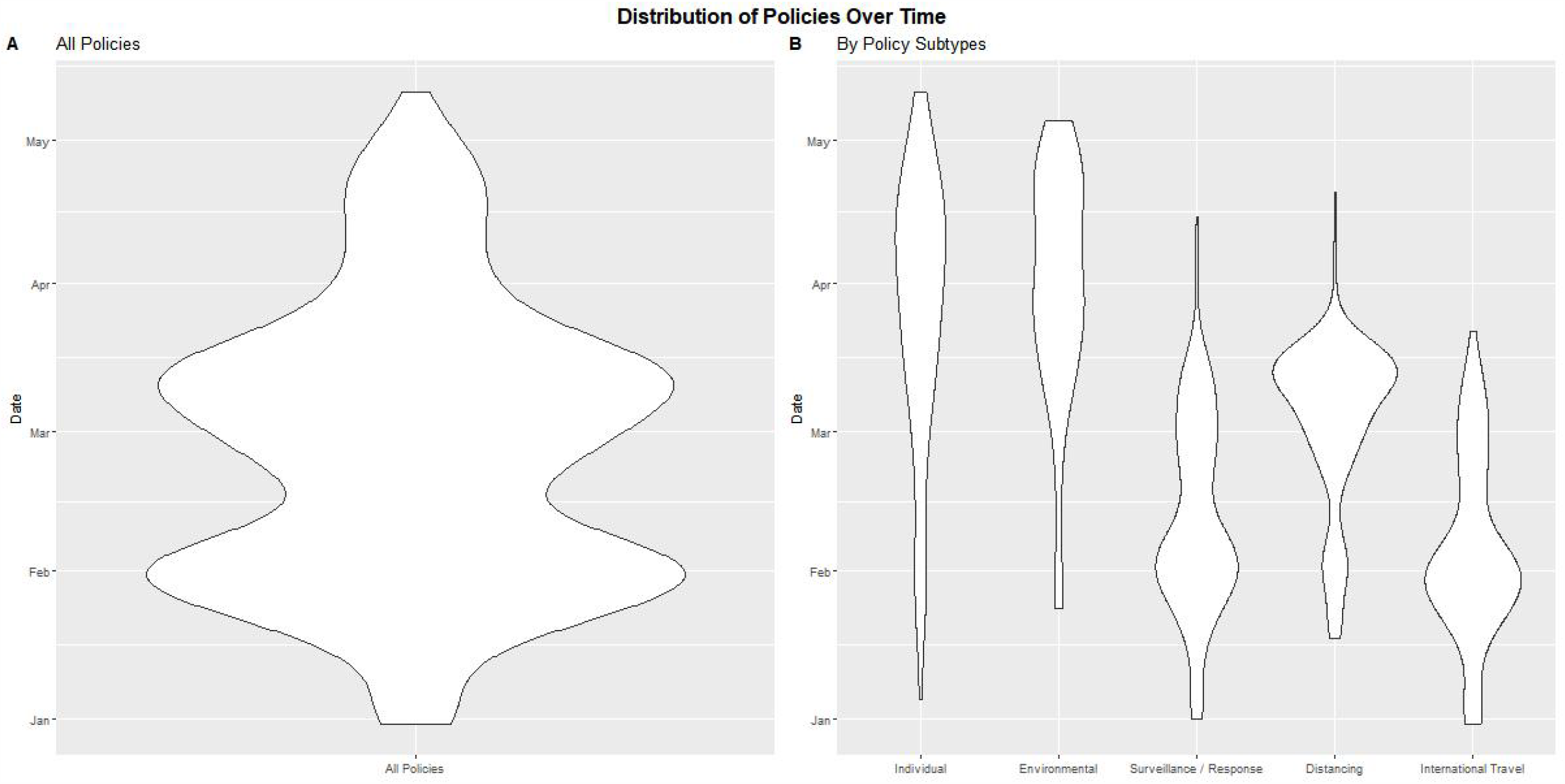
Distribution of policies over time

The relationship of the three independent variables are also plotted out graphically against the timeliness of any policy response for the dataset including H1N1 and specifying delay of response relative to Jan 5^th^, 2020. The effect any exposure to major epidemics showed the most striking negative relationship. The total number of cases and years since previous epidemics did not demonstrate much association with timeliness of response. This graphical relationship is demonstrated in figure 2, and is similar to the graphical representation that excluded H1N1 (available upon request).

**Figure 2:**
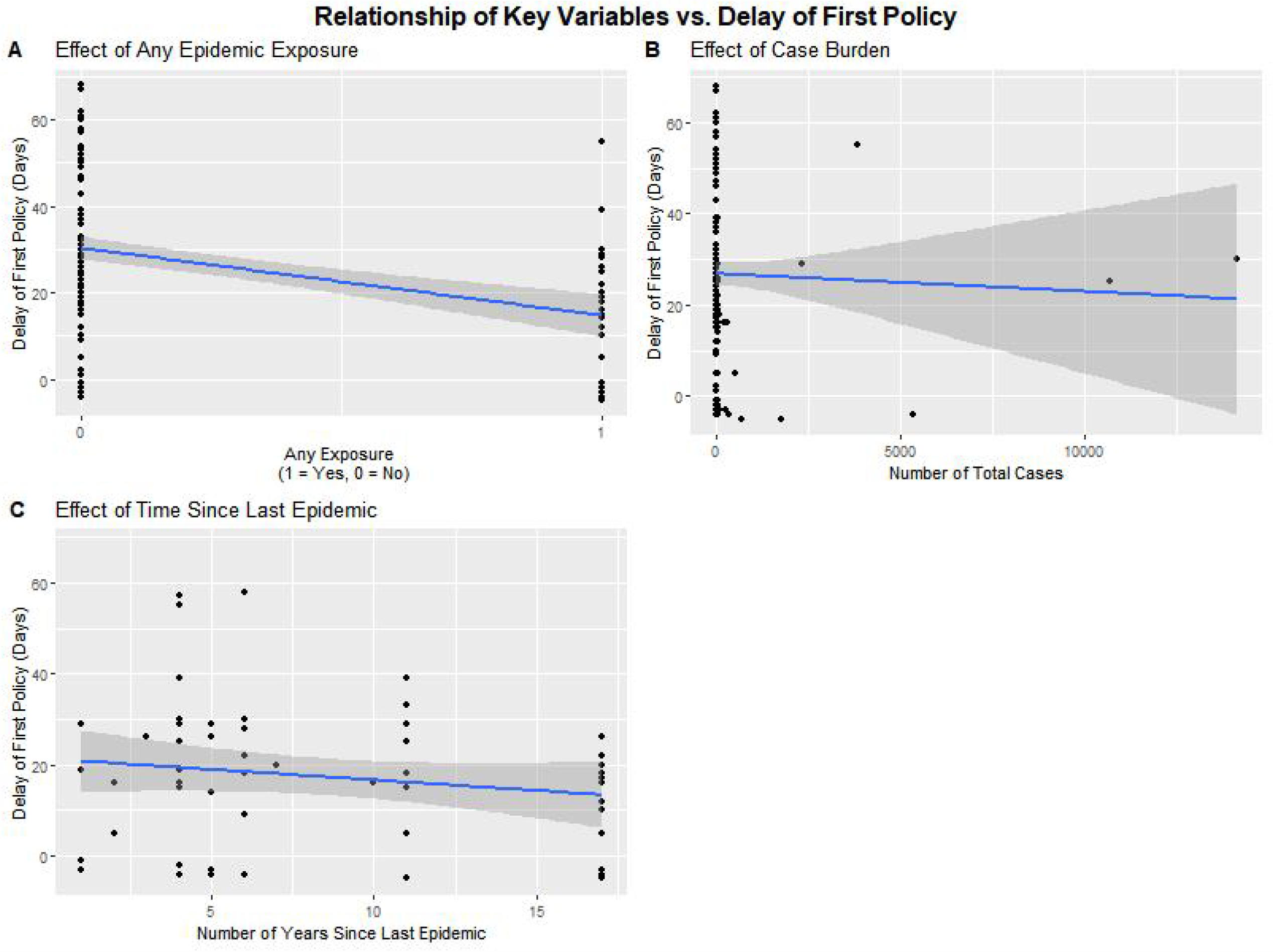
Bivariate relationship between key independent variables and delay of any first policy

### Main results

Results of analysis including H1N1 suggest that any epidemic exposure is associated with timelier policy responses when examining all policies, or within the categories of surveillance / response, distancing measures, and international travel policies by itself. Where significant, the effect of any epidemic exposure was to shorten the policy response by about six to ten days. Once the total cases are introduced, only the subcategories of policies demonstrated significant association between previous exposure and timeliness. The significance of this variable for any categories of policies is nullified when examined alongside years since previous epidemics.

The epidemics exposure was not significantly associated with timelier individual or environmental policies in any of the specifications. Part of the reason for the lack of significance may be due to the limited degrees of freedom available for the estimations in the individual or environmental policies, as well as for any specifications involving years since epidemics.

The total number of cases and years since the previous epidemics were not significant in any of the specifications. The results of all permutations of the regressions are provided in Table 4.

**Table 4:** Delay of timeliness relative to Jan 5, 2020

|  |  | Any policy | Individual | Environmental | Surveillance<br>/ Response | Distancing | International<br>Travel |
| --- | --- | --- | --- | --- | --- | --- | --- |
| Single<br>independent<br>variables | **Any Epidemics** | -8.0410<sup>*</sup><br>(3.6226) | -5.9035<br>(8.8169) | -10.3716<br>(7.4316) | -8.8626<sup>*</sup><br>(4.0429) | -6.0304<sup>*</sup><br>(2.8531) | -9.9082<sup>*</sup><br>(3.9583) |
|  | Adj. R<sup>2</sup> | 0.2705 | 0.2128 | 0.2461 | 0.3144 | 0.4909 | 0.2235 |
|  | Num. obs. | 148 | 69 | 78 | 147 | 147 | 148 |
|  | **Total Number of**<br>**Cases** | -0.0012<br>(0.0009) | 0.0031<br>(0.0046) | 0.0004<br>(0.0012) | -0.0008<br>(0.0010) | -0.0001<br>(0.0007) | -0.0011<br>(0.0010) |
|  | Adj. R<sup>2</sup> | 0.2548 | 0.2129 | 0.2253 | 0.2932 | 0.4743 | 0.1957 |
|  | Num. obs. | 148 | 69 | 78 | 147 | 147 | 148 |
|  | **Year Since Last**<br>**Epidemics** | 0.0241<br>(0.4000) | 0.2363<br>(1.4658) | -0.5234<br>(1.1168) | 0.1883<br>(0.4104) | -0.3189<br>(0.3280) | -0.0795<br>(0.4842) |
|  | Adj. R<sup>2</sup> | 0.2344 | -0.0454 | 0.1698 | 0.446 | 0.6478 | 0.1705 |
|  | Num. obs. | 52 | 27 | 31 | 52 | 52 | 52 |
| Two<br>independent<br>variables | **Any Epidemics** | -7.1323<br>(3.8315) | -7.4451<br>(9.0338) | -13.2051<br>(8.0233) | -8.7531<sup>*</sup><br>(4.2915) | -6.4996<sup>*</sup><br>(3.0211) | -9.3745<sup>*</sup><br>(4.1926) |
|  | **Total Number of Cases** | -0.0007 | 0.0039 | 0.0012 | -0.0001 | 0.0004 | -0.0004 |
|  |  | (0.0009) | (0.0047) | (0.0013) | (0.0010) | (0.0007) | (0.0010) |
| Adj. R<sup>2</sup> |  | 0.2681 | 0.2084 | 0.2448 | 0.3093 | 0.488 | 0.2187 |
| Num. obs. |  | 148 | 69 | 78 | 147 | 147 | 148 |
|  | **Any Epidemics** | -5.6692 | 7.1027 | -16.6054 | -3.1411 | -5.1064 | -10.918 |
|  |  | (5.1394) | (19.1462) | (16.3505) | (5.3304) | (4.2011) | (6.0704) |
|  | **Year Since Last Epidemics** | -0.1456 | 0.3776 | -1.3373 | 0.0943 | -0.4717 | -0.4063 |
|  |  | (0.4276) | (1.5571) | (1.3738) | (0.4435) | (0.3495) | (0.5050) |
| Adj. R<sup>2</sup> |  | 0.2385 | -0.1092 | 0.1712 | 0.4368 | 0.652 | 0.2143 |
| Num. obs. |  | 52 | 27 | 31 | 52 | 52 | 52 |
|  | **Total Number of Cases** | -0.0011 | 0.0026 | 0.0003 | -0.0005 | -0.0002 | -0.0008 |
|  |  | (0.0009) | (0.0077) | (0.0024) | (0.0010) | (0.0008) | (0.0011) |
|  | **Year Since Last Epidemics** | -0.0646 | 0.3412 | -0.4686 | 0.1506 | -0.3368 | -0.1404 |
|  |  | (0.4040) | (1.5437) | (1.2322) | (0.4211) | (0.3373) | (0.4954) |
| Adj. R<sup>2</sup> |  | 0.2440 | -0.1113 | 0.1244 | 0.4354 | 0.6396 | 0.1593 |
| Num. obs. |  | 52 | 27 | 31 | 52 | 52 | 52 |
| <u>All three independent</u> | **Any Epidemics** | -5.3146 | 6.3976 | -17.7499 | -2.9931 | -5.0487 | -10.699 |
|  |  | (5.1250) | (19.9700) | (17.1268) | (5.3945) | (4.2606) | (6.1317) |
| variables | **Total Number of Cases** | -0.0011<br>(0.0009) | 0.0022<br>(0.0080) | 0.0008<br>(0.0025) | -0.0005<br>(0.0010) | -0.0002<br>(0.0008) | -0.0007<br>(0.0011) |
|  | **Year Since Last Epidemics** | -0.2192<br>(0.4303) | 0.4552<br>(1.6349) | -1.2421<br>(1.4384) | 0.0635<br>(0.4529) | -0.4837<br>(0.3577) | -0.4517<br>(0.5148) |
|  | Adj. R<sup>2</sup> | 0.2454 | -0.1874 | 0.1280 | 0.4252 | 0.6433 | 0.2012 |
|  | Num. obs. | 52 | 27 | 31 | 52 | 52 | 52 |

**Note:** \*\*\*, \*\*, and \* indicate $p < 0.001$ , $0.01$ , and $0.05$ , respectively. The output from all other covariates are suppressed. Each observation reflects one single policy from a health system. An N of 52, for example, would suggest 52 health systems' first policies were analyzed.

The results without H1N1 are provided in Table 5. It showed somewhat similar results though there are even less occasions of statistical significance. The effect of any epidemics exposure is significant mainly when examined on its own for distancing and international travel measures, and when examined for environmental measures and distancing measures in estimation with total cases. Even when significant, the effects were only borderline significant.

**Table 5:**
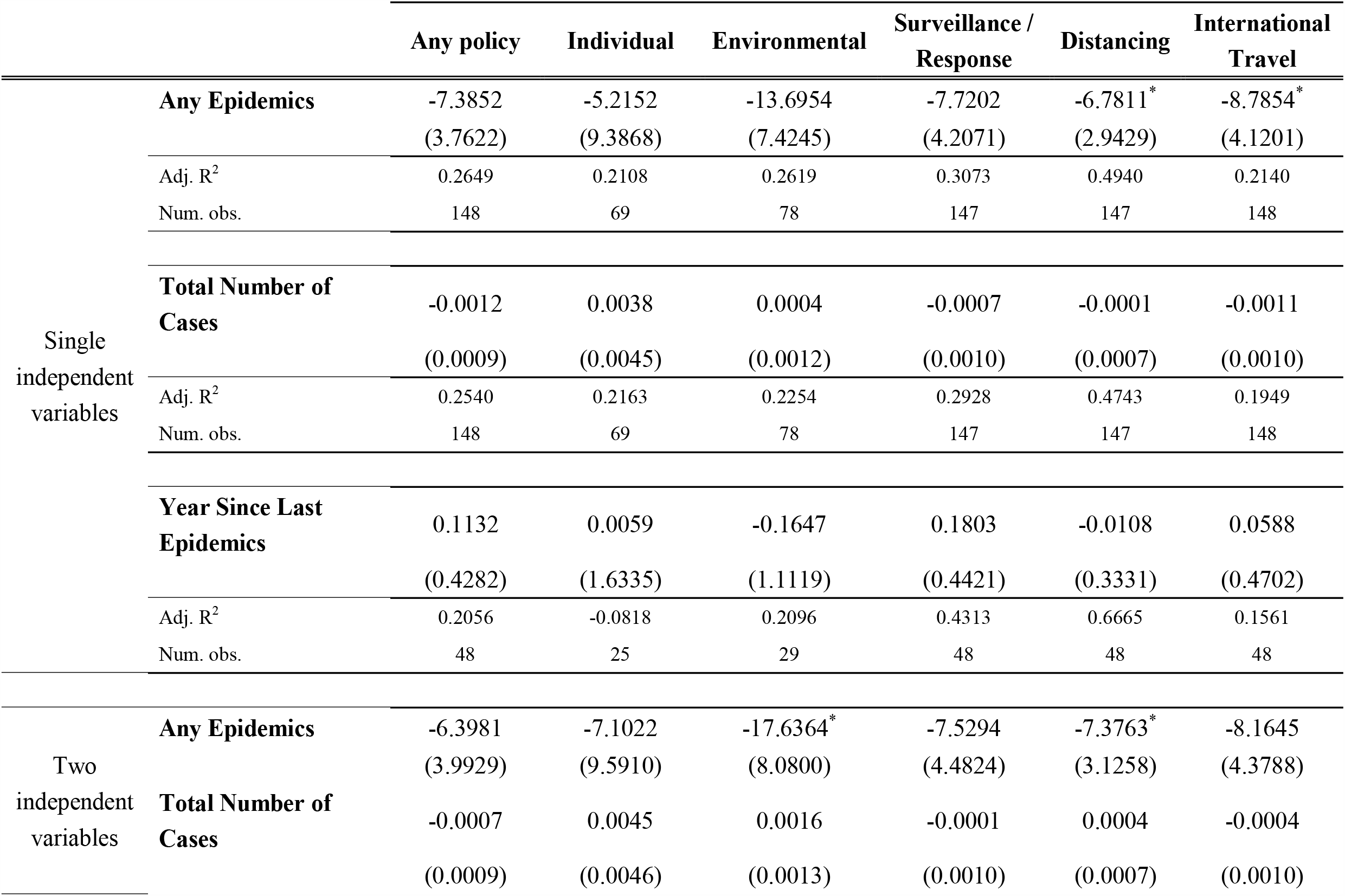
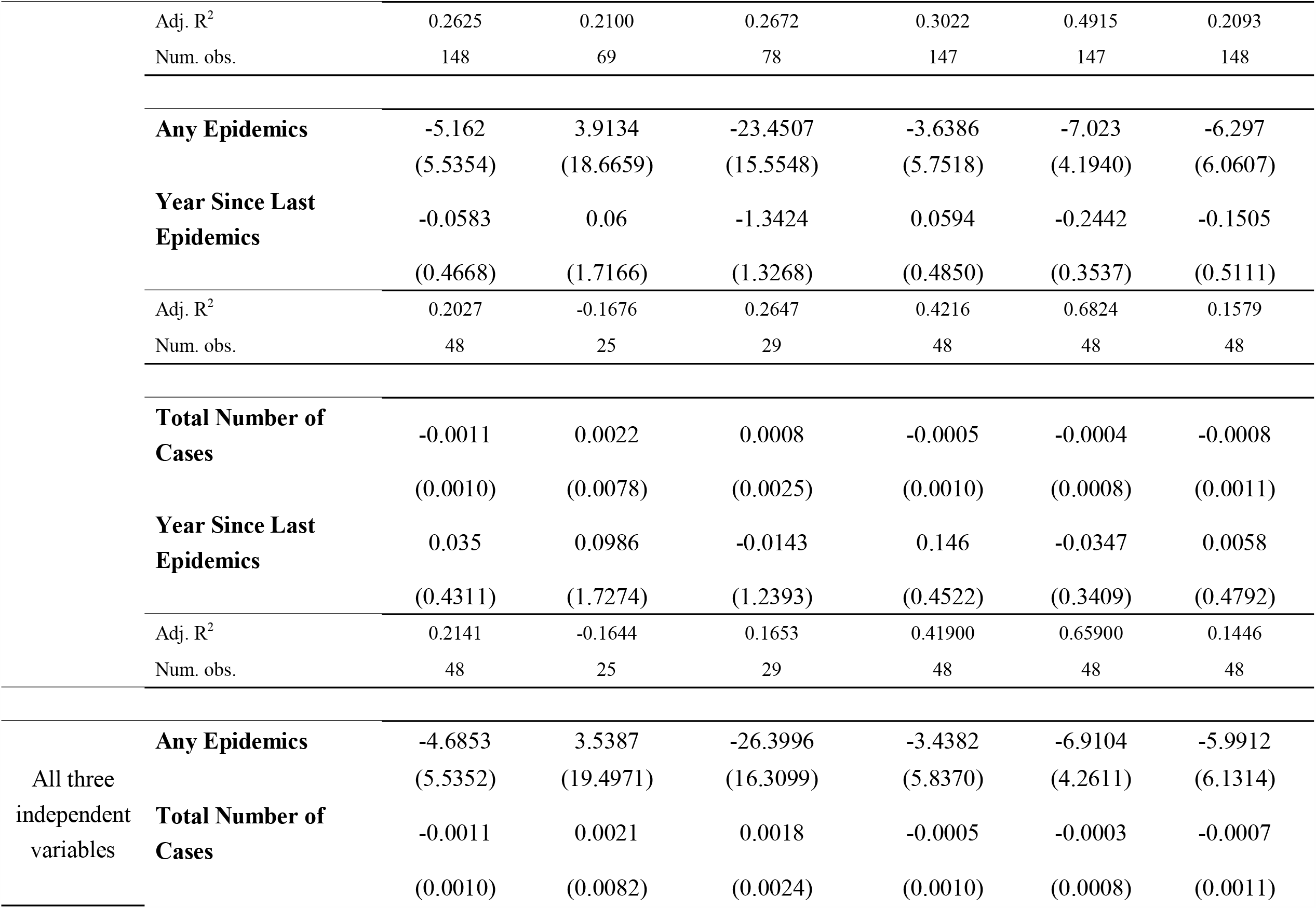
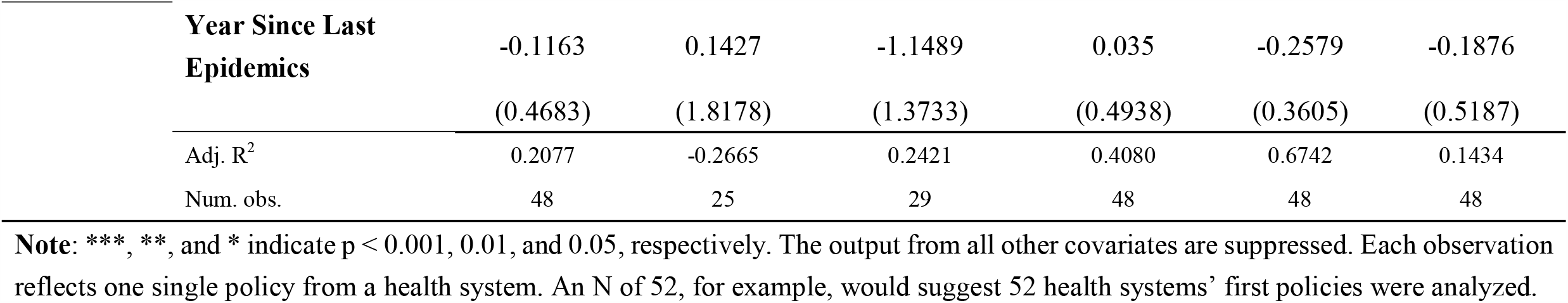
Regression results for dataset excluding H1N1

The results that specified policy delays relative to December 31^st^, 2019 shifted the intercepts by five. Since the parameters of interest are the coefficients and they remain unchanged, these additional results are not shown.

## Discussion

### Key results and interpretation

This study is, as far as I know, the first to formally apply organizational memory theory to explore the timeliness of COVID-19 response. This study identified that there may be shorter delays to initial policies when exposed to any previous epidemics within the last 20 years. The association between policy timeliness and organizational memory was significant only relative to any epidemics exposure, but not relative to the number of total cases or the recency of the exposures. The lack of significance for total cases or recency, however, may have been restricted by the sample size.

The policy response timing was quicker by about six to ten days. Relative to the average delay of policies that was about 53 days, the timeliness improved by about 11% to 19% faster among health systems with previous epidemics exposure. This seemed to be a moderate sized effect, and given the potential for exponential increase of infectious diseases, a respectably faster response. Developing correlations of faster response with burden of disease though would perhaps be useful next steps.

### Policy implication

Since this study showed that exposure to recent epidemics compared no such exposures was associated with faster response, it suggests that health systems do learn from past mistakes. Although there is insufficient evidence in this study to suggest that such learnings erode over time, institutionalizing the learnings appropriately through more permanent policies may nevertheless be useful. It may also be useful for health systems to conduct exercises intermittently to jog the organizational memory.

### Future research avenues

Peeri et al. suggest that health systems may have learned to communicate with the public more regarding hygiene and establish quarantine processes, but political leaders had not appreciated the importance of transparency or leveraged technology for more prompt surveillance methods [16]. The call towards such higher order considerations for more timely responses resonates with the need for further reflections for timelier responses.

Additional reflections and research may also draw from the work of Van Bavel et al., which highlighted a variety of social and behavioural mechanisms that could be useful for responding to COVID-19 [17]. Among the strategies highlighted, the risk communication, public persuasion, and misinformation denouncement are particularly relevant in times of crisis as they can be quickly implemented. Exploring how health systems have incorporated relevant lessons into their executive branch of government may help inform policymakers what lessons they ought to incorporate into their future health systems coming out of COVID-19. One particularly meaningful avenue is to clarify whether timelier response translates to improvement in lowering deaths or economic costs. As COVID-19 is yet incomplete, future studies might follow up on this front once COVID-19 resolves.

### Limitations

The cross-sectional design limits the interpretation to association, not causation. Further, the lack of health systems previously exposed to major epidemics precludes generalization across all systems. Health systems with previous epidemics may have developed more timely reporting procedures of policies to WHO, in which case timelier policy responses may actually be merely be an artifact of timelier reporting. The data is limited in that it cannot tease apart such nuances. Furthermore, the study’s interpretation of organizational memory suggests that it is individuals who retain such memory. A tighter study would have variables on individuals’ beliefs, knowledge, and adaptations for epidemic. The current dataset precluded such granular examinations.

## Data Availability

All datasets and the associated R script will be uploaded as supplement.

## Acknowledgements

I would like to thank the following individual for dynamic discussions that helped propel this project: Professor Winnie Chi-Man Yip, Dr. Annie Haakenstad, Dr. Der-Hwa Tsuei, Ms. Kuei-Chun Wu, and Dr. Amy Tsai. I would also like to thank the two reviewers for their insightful and helpful comments, which significantly strengthened the quality of the work.

## Footnotes

1 This recoding mainly affected Hong Kong and Taiwan.

2 The *firstpol* variable was also specified relative to December 31^st^, 2019, but this changed only the intercept. This phenomenon is addressed further in the results section. The study therefore focused on presenting analysis using delay of policy relative to Jan 5, 2020.

3 For completeness, the variable was also defined relative to December 31, 2019, which was when China reported pneumonia of unknown significance to the WHO.

4 The WHO regions are defined as follows: AFRO, African region; AMRO, Americas; EMRO, Eastern Mediterranean; EURO, European Region; SEARO, South-East Asia Region; and WPRO, Western Pacific Region.

